# Impact of diabetes mellitus on the stabilization and osseointegration of dental implants: a retrospective study in Greece

**DOI:** 10.1101/2022.03.19.22272649

**Authors:** Aglaia Katsiroumpa, Petros Galanis, Ioannis Diamantis, Sofia Georgikopoulou, Theodoros Katsoulas, Olympia Konstantakopoulou, Elissavet Katsimperi, Evangelos Konstantinou

**Affiliations:** Faculty of Nursing, National and Kapodistrian University of Athens, Athens, Greece; Clinical Epidemiology Laboratory, Faculty of Nursing, National and Kapodistrian University of Athens, Athens, Greece; Athens, Greece; Center for Health Services Management and Evaluation, Faculty of Nursing, National & Kapodistrian University of Athens, Athens, Greece; School of Dentistry, National and Kapodistrian University of Athens, Athens, Greece

**Keywords:** diabetes mellitus, dental implant, osseointegration, stabilization

## Abstract

**Introduction:** Adequate dental restoration including the use of implants is critical in healthy eating habits of diabetic patients and appropriate metabolic control.

**Aim:** To investigate the relationship between diabetes mellitus and dental implants stabilization and osseointegration.

**Methods:** A retrospective study was conducted in a private dental clinic in Athens. Data collection referred to the period between January 2016 and August 2021. During this time period, all cases related to implant placement in diabetic patients at the clinic were recorded. In particular, 93 implants were recorded in 36 diabetic patients. During the same time period, 93 implant cases involving non-diabetics at the clinic were randomly taken from the clinic records to provide the comparison group. The implant stability quotient was measured immediately after implant placement and after four months.

**Results:** The mean value of the implant stability quotient immediately after implant placement was 75.97 in non-diabetics and 76.85 in diabetics (p=0.42). The mean value of the implant stability quotient after four months was 78.92 in non-diabetics and 78.44 in diabetics (p=0.58). The mean value of the implant stability quotient in non-diabetics increased statistically significantly in the first four months from 75.97 to 78.92 (p<0.001). The mean value of the implant stability quotient in diabetics increased statistically significantly in the first four months from 76.85 to 78.44 (p=0.011). No implant loss was recorded in both diabetics and non-diabetics (p=1). According to multivariate analysis, patients who did not have bio-materials placed during implantation, patients who had not undergone previous surgical procedures and patients who had implants placed in the mandible had better implant stability.

**Conclusions:** The stability of the implants increased statistically significant in the first four months of implant placement. No relationship was found between diabetes mellitus and dental implants stabilization and osseointegration. However, studies with a larger sample size and longer follow-up of patients are needed to better clarify the risks and benefits of dental implants in diabetic patients.

## Introduction

Diabetes mellitus is a metabolic disorder which leads to hyperglycaemia and therefore to vascular diseases. People with diabetes mellitus tend to have periodontal diseases, teeth losses, delay in healing and worse outcomes in infection diseases (Abiko & Selimovic, 2010). The prevalence of diabetes is constantly increasing, especially in developed countries (Danaei et al., 2011). For example, in 1980, more than 150 million people worldwide suffered from diabetes, while in 2008, this number exceeded 350 million people.

Dental implants are a successful treatment for replacing missing teeth, since the ten years survival rate of dental implants was reported to be 94.6% (Moraschini et al., 2015). Effective osseointegration process during healing period affects implant success (Fiorellini & Nevins, 2000). Moreover, the amount of osseointegration is affected by several risk factors including smoking, radiotherapy, osteoporosis, diabetes, etc. (Chen et al., 2013). Diabetic patients have an increased incidence of periodontitis and tooth loss, delayed wound healing and worse infection outcomes (Abiko & Selimovic, 2010; Khader et al., 2006).

The role of implants in the case of diabetic patients is extremely important, as after tooth loss these patients avoid foods that cause them difficulty in chewing, resulting in a poor diet. Adequate dental rehabilitation with the use of implants is essential in promoting the eating habits of diabetic patients and better metabolic control (Chrcanovic et al., 2014). Several systematic reviews have investigated the effect of diabetes on the stabilization and osseointegration of dental implants resulting on mixed results (Andrade et al., 2021; Chen et al., 2013; Chrcanovic et al., 2014; Katsiroumpa et al., 2022; Oates et al., 2013; Shang & Gao, 2021). In general, when diabetes is under control, implant procedures are safe and diabetic patients seem to be able to achieve a survival rate of dental implants like that of non-diabetics.

The aim of this study was to investigate the relationship between diabetes mellitus and dental implants stabilization and osseointegration.

## Methods

### Study design

We conducted a retrospective study in a dental clinic in Athens. Data collection time was from January 2016 to August 2021. All the cases of dental implantations in patients with diabetes mellitus were registered during that period. More specific, there were 93 implants in 36 patients with diabetes mellitus. Simultaneously, there were randomly chosen 93 implants from the archives of the clinic, concerning non diabetic patients which comprised the control group. The implants that referred to the control group were occurred from a random table number. The study outcomes were the resonance frequency analysis and the loss of the dental implant.

The resonance frequency of the implants is calculated by the analysis of the resonance of the frequency. Practically, that means the calculation of the stabilization quotient of the implant, and takes numbers from 1 to 100. Higher values indicate higher stabilization. The resonance frequency analysis was performed immediately after the implantation and, again, after four months so that the stabilization and the osseointegration of the dental implant could be investigated. The four-months-time period was determined due to the fact that two weeks after the implantation is the minimum stabilization which can be observed and then follows the phase of osseointegration which takes three to four weeks.

The loss of the dental implant is determined by the following parameters: (a) the stabilization quotient of the implant, (b) the x-ray which depicts the implant situation, and (c) the clinical picture and examination.

The information concerning the diabetes mellitus was occurred from the medical history of the patients. Furthermore, we recorded potential confounders, in order to eliminate them with the multivariate analysis. We recorded the following confounders: sex, age, smoking, medication for chronic disease, cardiovascular diseases, respiratory diseases, cancer, high blood pressure, autoimmune diseases, thyroid diseases, diseases of the digestive system, infectious diseases, previous surgical operations, allergies in medication, use of biomaterial for the implantation and immediate implantation after the tooth extraction.

### Ethical issues

Data were collected from the clinic archives after the written permission of the scientific directors of the clinic. We did not collect personal data of the patients. We obtained written informed consent of patients. Study protocol was approved by the Faculty of Nursing, National and Kapodistrian University of Athens (reference number: 385, date 14/1/2022).

### Statistic analysis

We use frequencies (percentages) to present categorical variables and mean (standard deviation) to present continuous variables. According to the Kolmogorov-Smirnov test and normal Q-Q plots, continuous variables followed normal distribution. Bivariate analysis between independent and dependent variables included chi-square test, independent samples t-test, paired samples t-test, analysis of variance, Pearson’s correlation coefficient and Spearman’s correlation coefficient. We performed multivariate linear regression with the stabilization quotient of the implants as the dependent variable. In that case, we present adjusted coefficients beta, 95% confidence intervals (CIs) and p-values. All tests of statistical significance were two-tailed. Statistical analysis was performed with the Statistical Package for Social Sciences software (IBM Corp. Released 2012. IBM SPSS Statistics for Windows, Version 21.0. Armonk, NY: IBM Corp.).

## Results

### Demographic and clinical characteristics

The 93 implants involving diabetic patients placed in 36 patients, while the 93 implants involving non-diabetic patients placed in 42 individuals. Among participants, 52.6% (n=41) were females and 47.4% (n=37) were males. The mean age of the participants was 57.6 years (standard deviation was 11.7) and 23.1% (n=18) were smokers.

More than half of the participants (56.4%) were taking medication for a chronic disease, while 21% had undergone previous surgery and 2.2% had drug allergies. The most common diseases among the participants were cardiovascular diseases (43.6%), hypertension (37.2%), thyroid gland diseases (7.7%) and digestive system diseases (6.4%).

Fifty point five percent of the implants were placed in the mandible and 49.5% in the maxilla. In 16.1% of cases, the implant was placed directly, while in 8.6% of cases, bio-materials were placed during implant placement.

Fifty point five percent of the implants were placed in the mandible in both diabetics and non-diabetics (p=0.99). Placement of bio-materials during implantation was more frequent in non-diabetics (17.2% vs. 0%, p<0.001). In 18.3% of cases in non-diabetics, implant placement was immediate, while the corresponding percentage in diabetics was 14% without this difference being statistically significant (p=0.43).

### Stabilization quotient and loss of the implants according to diabetic status

Mean stabilization quotient of the implants immediately after implant placement was 75.97 in non-diabetics and 76.85 in diabetics without this difference being statistically significant (p=0.42). Also, mean stabilization quotient of the implants after four months was 78.92 in non-diabetics and 78.44 in diabetics without this difference being statistically significant (p=0.58).

Mean stabilization quotient of the implants in non-diabetics increased statistically significant in the first four months from 75.97 to 78.92 (p<0.001). Moreover, mean stabilization quotient of the implants in diabetics increased statistically significantly in the first four months from 76.85 to 78.44 (p=0.011).

No implant loss was recorded in both diabetics and non-diabetics (p=1).

### Factors related with stabilization quotient of the implants

Bivariate analysis between independent variables and stabilization quotient are shown in Table 1. Then we performed multivariate linear regression with the stabilization quotient of the implants as the dependent variable and we found that patients who did not have bio-materials placed during implantation (coefficient beta=4.41, 95% CI=0.55 to 8.27, p=0.025), patients who had not undergone previous surgical procedures (coefficient beta=2.86, 95% CI=0.44 to 5.29, p=0.021) and patients who had implants placed in the mandible (coefficient beta=4.25, 95% CI=2.24 to 6.25, p<0.001) had better implant stability (Table 2).

**Table 1.**
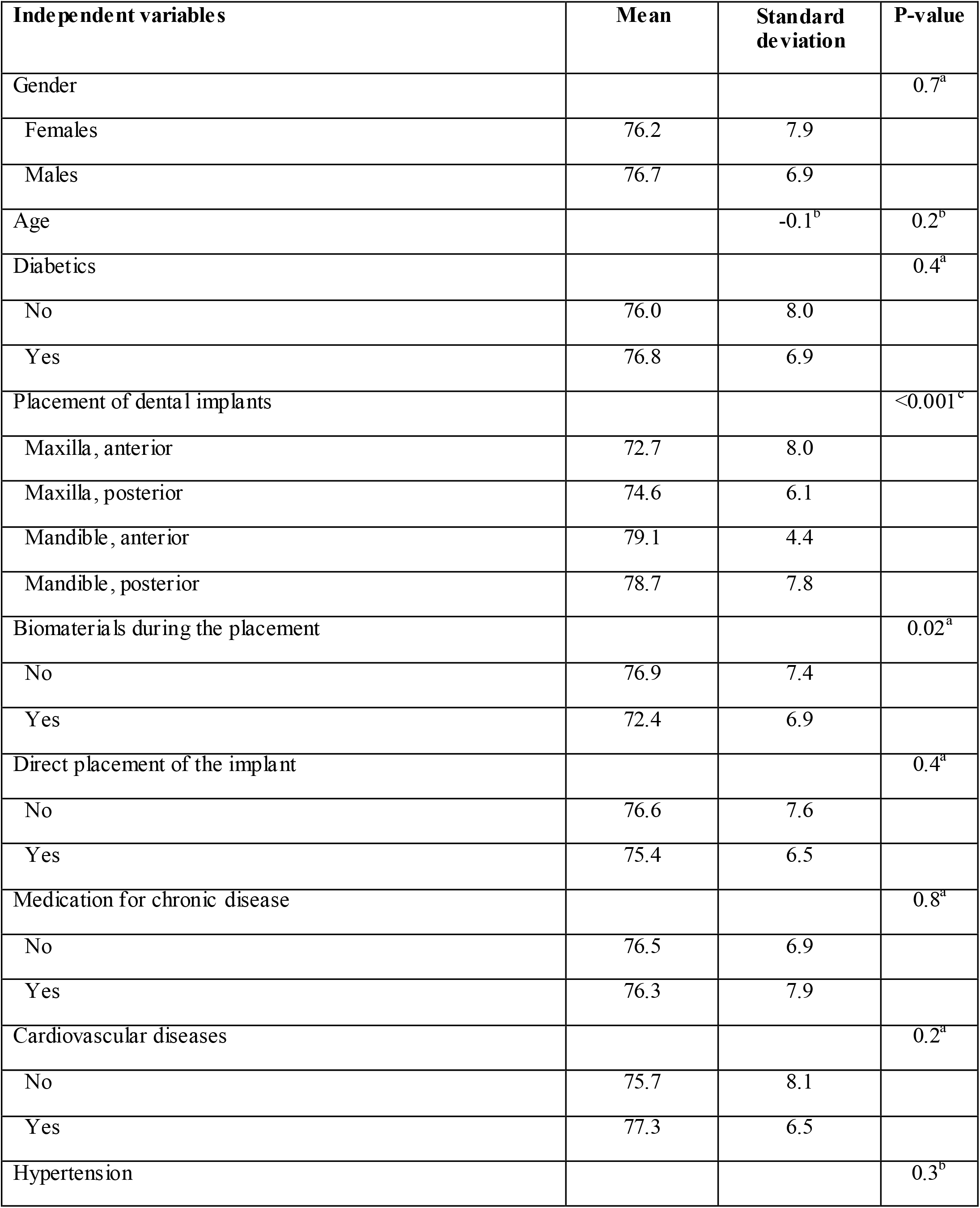
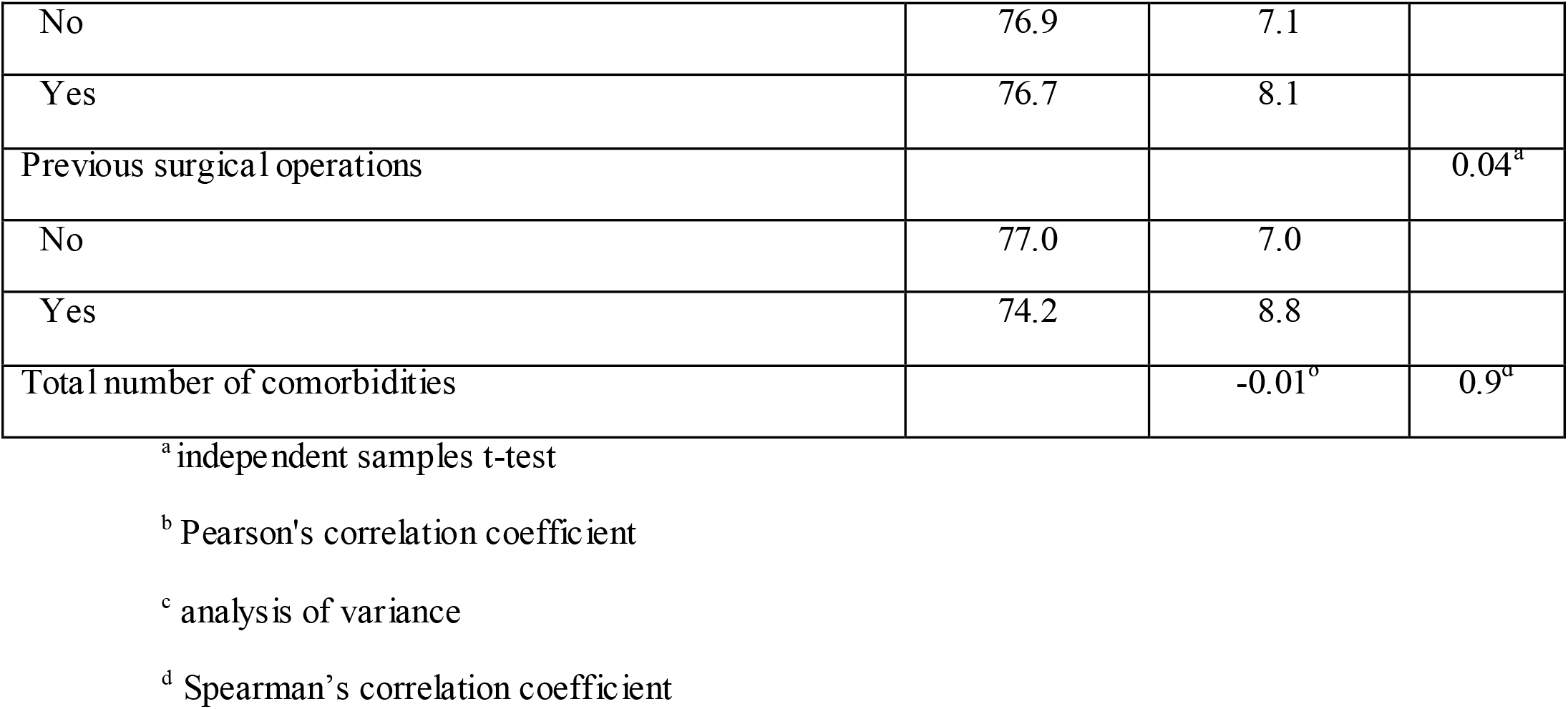
Bivariate analysis between independent variables and stabilization quotient.

**Table 2.** Multivariate linear regression with the stabilization quotient of the implants as the dependent variable.

| Independent variable | Coefficient<br>beta | 95% confidence interval<br>for beta | P-value |
| --- | --- | --- | --- |
| Implants placed in the mandible vs. implants placed in the maxilla | 4.25 | 2.24 έως 6.25 | <0.001 |
| Patients who did not have bio-materials placed during implantation vs. patients who had bio-materials | 4.41 | 0.55 έως 8.27 | 0.025 |
| Patients who had not undergone previous surgical procedures vs. patients who had undergone surgical procedures | 2.86 | 0.44 έως 5.29 | 0.021 |

## Discussion

A retrospective study was performed by collecting data from a dental clinic in Athens to investigate the relationship of diabetes mellitus with the stabilization and osseointegration of dental implants. The study included 93 implants placed in diabetic patients and 93 implants placed in non-diabetic patients as the control group.

Mean stabilization quotient of the implants in the first four months increased statistically significant in both diabetics and non-diabetics. Specifically, mean stabilization quotient of the implants in diabetics increased from 76.85 to 78.44, while in non-diabetics increased from 75.97 to 78.92. These findings are confirmed by the literature, as in three studies, mean stabilization quotient of the implants increased statistically significant in healthy subjects (Ghiraldini et al., 2016; Oates et al., 2014; Sundar et al., 2019), while in three studies, mean stabilization quotient of the implants increased statistically significant in diabetics (Al Zahrani & Al Mutairi, 2018; Oates et al., 2014; Sundar et al., 2019). Mean stabilization quotient of the implants shows high variability among different studies which may be due to various parameters such as different study populations, different types of diabetes, different level of diabetes control, etc. For instance, in the study by Ghiraldini et al. (2016), mean stabilization quotient of the implants after implant placement was 79.36 in non-diabetics, 79.77 in diabetics with poor diabetes control and 80.17 in diabetics with good diabetes control, while three months after implant placement, the respective values were 80.11, 78.33 and 80.13. However, in a similar study by Oates et al. (2014), the values of mean stabilization quotient of the implants were significantly lower and more specifically, mean stabilization quotient of the implants immediately after implant placement was 58 in non-diabetics and 63.8 in diabetics, while four months after implant placement, the respective values were 62.2 and 65.6.

In addition, in the present study, all participants retained their implants. The same conclusion was reached by seven studies in which all diabetics and non-diabetics were found to retain their implants during the study (Al Amri et al., 2016; Alsahhaf et al., 2019; Al-Shibani et al., 2019; Dowell et al., 2007; Erdogan et al., 2015; Gómez-Moreno et al., 2015; Sundar et al., 2019). Regarding implant loss, four studies found statistically significant more frequent implant loss in diabetics (Daubert et al., 2015; Loo et al., 2009; Moy et al., 2005; Zupnik et al., 2011), while five studies found that implant loss was more frequent in diabetics but was not statistically significant (Aguilar-Salvatierra et al., 2016; Morris et al., 2000; Ormianer et al., 2018; Sghaireen et al., 2020; Tawil et al., 2008). In contrast, ten studies found that implant loss was more frequent in healthy subjects but was not statistically significant (Alsaadi et al., 2008; Anner et al., 2010; Bell et al., 2011; Busenlechner et al., 2014; Doyle et al., 2007; Keller et al., 1999; Le et al., 2013; Levin et al., 2011; Oates et al., 2014; van Steenberghe et al., 2002).

According to multivariate analysis, there was no association between diabetes mellitus and implant stabilization quotient. This finding is confirmed in six studies in which the analysis of implant resonance frequency was performed and no statistically significant difference was found between healthy and diabetic patients (Al Zahrani & Al Mutairi, 2018; Erdogan et al., 2015; Ghiraldini et al., 2016; Oates et al., 2009; Sundar et al., 2019). The lack of difference between diabetic and non-diabetic patients may be due to the fact that diabetic patients were under control, resulting in extremely limited negative effects of the disease on osseointegration. This fact is supported by studies in which it was found that good diabetes control and reduction of hyperglycaemia led to better faster fracture repair (Beam et al., 2002; Funk et al., 2000; Gebauer et al., 2002). The exact opposite is also true in patients with poorly controlled diabetes, with hyperglycaemia causing vascular complications and reducing the ability to heal (Amir et al., 2002; Cohen & Horton, 2007; Funk et al., 2000; Lu et al., 2003; Okazaki et al., 1999; Vieira Ribeiro et al., 2011). It should be noted that the absence of a relationship between diabetes mellitus and stabilization quotient of implants is confirmed by studies that lasted up to one year after implant placement. In the present study, the time limit of four months was chosen, as two to four weeks after implant placement, the minimum stabilization quotient is recorded, followed by the process of osseointegration, which takes approximately three to four months (Ghiraldini et al., 2016; Oates et al., 2014).

The present study found that patients in whom no bio-materials were placed during implantation, patients who had not undergone previous surgical procedures and patients in whom implants were placed in the mandible had better implant stability. These findings are reasonable, as bio-materials are placed in patients with greater bone loss which is associated with worse oral cavity condition, so as to enhance the ability of dental implants to osseointegration and increase their stability. In addition, patients who have not undergone previous surgical procedures are in better physical condition and have less morbidity which favors better bone metabolism and consequently better osseointegration of dental implants. The denser bone composition in the mandible compared to the maxilla could explain the fact that patients who received implants in the mandible had better implant stability.

## Limitations

This study also had some limitations. Specifically, participants were separated into diabetic and non-diabetic patients after self-reporting during the history taking in the clinic. No glycemic control of the participants was performed which introduces information bias in the study as there is a possibility that some non-diabetics are under-diagnosed. In addition, the inability to perform glycemic control did not enable the diabetic patients to be distinguished into those with good diabetes control and those with poor control. A similar information bias may be introduced into the study by other information recorded from the history of the patients attending the clinic, e.g. smoking habit, coexisting conditions, etc. In addition, confounders present in the patients’ history were recorded, as data collection was performed retrospectively based on the record of patients registered at the clinic. Therefore, there may be other confounders that were not measured in the study as they were not included in the patients’ history. In addition, the type of diabetes, i.e. whether they had type I or type II diabetes, was not recorded in the patients’ history. In addition, measurements of stabilization quotient were performed immediately after implant placement and after four months. Following the patients for a longer period of time could provide more information about the stabilization and osseointegration of the dental implants.

## Conclusions

In the present study, no relationship was found between diabetes mellitus and stabilization quotient of the implants. This study contributed information to this field of research, but the effect of diabetes and glycemic control on the stabilization and osseointegration of dental implants requires further studies with better design and less error to draw safer conclusions. For this reason, studies with a larger sample size and longer follow-up of patients are needed to better clarify the risks and benefits of dental implant placement in diabetic patients.

## Data Availability

All data produced in the present study are available upon reasonable request to the authors

